# Methods of An Open-Label Proof-Of-Concept Trial of Intravenous Valproic Acid for Severe COVID-19

**DOI:** 10.1101/2020.04.26.20079988

**Authors:** Erwin Chiquete, Liz Toapanta-Yanchapaxi, Carlos Cantú-Brito

## Abstract

**Introduction:** Coronavirus disease 2019 (COVID-19) is the systemic entity caused by the severe acute respiratory syndrome coronavirus 2 (SARS-CoV-2) that may cause death through severe atypical pneumonia and acute lung injury. Valproic acid (VPA) has shown anti-inflammatory activity and mild intrinsic antiviral effect. These properties warrant the study of VPA as a possible active treatment in persons with severe COVID-19.

**Methods:** Consecutive adult patients needing invasive mechanical ventilation (IMV) will be given intravenous (i.v.) VPA at a starting dose of 20 mg/kg/day and up to 60/kg/day (in 60 min i.v. infusions in 250 mL normal saline) as needed to reach plasma VPA concentrations of 50–100 μg/mL (measured every 72 h). These patients will be followed-up for 10 days for the primary outcome and for a further period of 30 days after treatment completion for the secondary outcome of recurrence. The primary study outcome is the reduction in the case fatality rate of at least 50% after 10 days of treatment (as compared with natural history). Secondary outcomes are the reduction of length of stay (LOS) of at least 50%, as well as COVID-19 recurrence at 30-day follow-up. The most important safety outcomes are acute liver failure, acute pancreatitis, and thrombocytopenia.

**Conclusion:** Although long-term adverse effects and even pro-inflammatory consequences have been reported with the chronic use of VPA, given the urgent need for a drug against COVID-19 to shorten the high mortality and LOS, the study of VPA is justified from a scientific standpoint.

## Introduction

Coronavirus disease 2019 (COVID-19) is the systemic disease caused by the severe acute respiratory syndrome coronavirus 2 (SARS-CoV-2) that may cause death through severe atypical pneumonia and acute lung injury. There is a need for useful molecules that may cut the rate of deaths and hospital length of stay (LOS) that produce such a high sanitary burden. Given the urgent need for high-efficacy drugs, repurposed molecules may be the most efficient approach given the scarce time to limit the sanitary and economic impact of the present pandemics.

Valproic acid (VPA, the conjugate base is valproate) is available as sodium, semi sodium, and magnesium. VPA is a drug primarily used to treat epilepsy, bipolar disorder, as well as migraine.^1^ Chemically VPA is a weak organic acid that was first synthesized in 1882 as an analog of valeric acid, found naturally in the plant *valeriana officinalis*.^1,2^ VPA was tested initially as a solvent of organic compounds, and latter found serendipitously to be active in animals and humans as an anticonvulsant in 1962.^3^ It is a carboxylic acid clear liquid at room temperature. VPA was first approved in France (1967) for use in humans as an anti-epileptic medication.^1^

VPA attenuates the expression of pro-inflammatory cytokines in preclinical models^4–9^ and in humans^10,11^ especially during the acute treatment phase, although conflicting results have been reported.^12^ This molecule has also shown intrinsic antiviral activity probably due to its activity as histone deacetylase inhibitor.^9,13–16^ VPA has shown anti-inflammatory activity in myocardial^17^ as well as neural tissue in models of brain and spinal cord injuries,^18,19^ although with some conflicting findings.^20^

Although long-term adverse effects and even pro-inflammatory consequences have been reported with the chronic use of VPA,^12^ we believe that, given the urgent need to have a useful drug to shorten the high mortality and LOS associated with COVID-19,^21,22^ the study of the safety and efficacy of VPA in the treatment of COVID-19 is warranted from a scientific standpoint.

## Methods

### Study type

Open-label proof-of-concept interventional study.

### Hypothesis

Intravenous valproic acid will be safe and effective in the treatment of patients with severe COVID-19.

### Intervention

Sodium valproic acid will be given intravenously at a starting dose of 20 mg/kg/day and up to 60/kg/day as needed to reach plasma valproic acid concentration of 50–100 μg/mL (measured every 72 h). Three equal doses per day given intravenously in infusions lasting 60 min in 250 mL normal saline.

### Study oversight and ethical considerations

Ten days of i.v. treatment for the primary outcome, plus 30 days for the secondary outcome of COVID-19 recurrence, given a total of 40 days of follow-up.

### Population

Patients admitted with severe COVID-19 to the ICU of the *Instituto Nacional de Ciencias Médicas y Nutrición Salvador Zubirán* (INCMNSZ).

### Main study outcomes

Primary outcomes:

- Reduction in a case fatality rate (CFR) of at least 50%, from a known proportion of 40% to 20% after 10 days of treatment.

Secondary exploratory outcomes:

- Reduction of length of stay (LOS) of at least 50%, from a known median LOS of 10 days to 5 days after at least 5 days of i.v. valproic acid exposition.
- No COVID-19 recurrence during a 30-day observational period, after completion of the 10-day i.v. therapy.

### Selection criteria

Inclusion criteria:

- Patients aged >18 years
- Both sexes
- Positive to SARS-CoV-2 RNA in nasopharyngeal samples by RT-PCR
- Severe acute respiratory syndrome treated with IMV at study recruitment
- Any time elapsed since symptoms onset to study recruitment
- Any length of hospital stay
- Any length of IMV
- Signed informed consent from patient or legal proxy
- No known VPA allergy
- Patients with or without previous or current exposure to anti-epileptics
- Patients with or without previous or current exposure to chloroquine, hydroxychloroquine or azythromicin
- Patients not participating in an interventional trial
- Patients not under current valproate treatment
- Patients not under current treatment with remdesivir
- Patients not under current treatment with tocilizumab

Exclusion criteria:

- Decline of informed consent
- Signs of VPA allergy
- Any acute severe side effect (either thrombocytopenia, acute liver disease, moderate-to-severe hypertransaminasemia, or acute pancreatic injury)
- Incomplete information for primary outcome

*Intention-to-treat analysis (ITT) will be performed, considering study dropouts as failures.

### Recruitment strategy

Consecutive patients needing IMV and meeting selection criteria will be given i.v. valproic acid for 10 days maintaining a plasma concentration of 50–100 μg/mL (measured every 72 h). These patients will be followed-up for 10 days for the primary outcome and for a further period of 30 days after treatment completion.

### Safety outcomes

The most important acute side effects of valproic acid that should be meticulously on a daily basis examined are:

- Acute liver failure, defined as the severe acute liver injury with encephalopathy and impaired synthetic function (international normalized ratio ≥1.5) in a subject free of preexistent chronic liver disease.
- Acute pancreatitis, defined as the American Gastroenterological Association (AGA)^23^ guidelines: the presence of two out of three of the following characteristics: a) abdominal pain consistent with the disease, b) serum amylase and/or lipase activity >3 times the upper limit of normal, and c) characteristic findings on abdominal imaging.
- Thrombocytopenia, defined as the platelet blood count <50,000 per μL.
- Hypertransaminasemia, defined as the elevation of alanine transaminase (ALT) and/or aspartate transaminase (AST) activity in serum, >3 times the upper normal limit of the laboratory reference.

As stated in the selection criteria, patients with the current use of anti-epileptics are allowed to be recruited, criteria that may include patients with epilepsy. In case of the need of other anti-epileptics to treat new or recurrent seizures, the inclusion of such medications will be a prerogative of the treating physician team. The investigational team of this trial will reinforce the maintenance of the patient in the study, by helping in the selection of anti-epileptics with no known adverse interactions with VPA, but in any particular case of perceived harm that may arise with the interaction of VPA with other anti-epileptics, the patient will be removed from the study.

### Study duration

Ten days of i.v. treatment for the primary outcome, plus 30 days for the secondary outcome of COVID-19 recurrence, given a total of 40 days of follow-up.

### Major statistical hypothesis

There will be at least a 50% reduction of death of patients with severe COVID-19 needing invasive mechanical ventilation (IMV).

### Sample size

This is a proof-of-concept study, therefore, sample size calculation was performed by using the formula for the difference of proportions with a known (previous) proportion, and assuming a high magnitude of the efficacy of the study drug, since, given the contingency of this pandemics, minor effect sizes reductions are not desirable.

According to the most extensive information available today on 1,591 Italian patients with severe COVID-19 admitted to ICU,^24^ it is expected a 40% CFR final disposition of patients, according to Bayesian analysis (the original information reports 26% CFR, but 58% of patients still in ICU as of the analysis deadline of March 25, 2020). Hence, the sample size was calculated to detect a minimum of (but not restricted to) 50% reduction in the CFR from 40% to 20%, with 80% study power and 5% type I error.

Therefore, a sample size of at least (but not restricted to) 43 patients will be needed to demonstrate an absolute (not RRR) reduction of 50% in CFR. No excess sample size was considered to cover study losses since ITT analysis requires that drop-outs are considered as failures to achieve study outcomes.

### Statistical plan

ITT analyses will be performed considering all patients removed from the study for any reason as treatment failures. Per protocol (PP) analyses will also be performed considering in the calculations only patients with complete information and complete treatment schemes, but the analyses on the PP population will only be considered as hypothesis-generating to further pursue the study objectives with another robust design. Daily patients inspections will be performed in order to demonstrate first that valproic acid is not harmful, to review plasma levels (measured every 72 h), and clinical response. With this information, on a daily basis, statistical analyses will be performed for outcome differences by using chi-squared tests for categorical variables and hazard ratios to obtain effect sizes of study outcomes (primary and secondary outcomes), as well as safety outcomes. After completion of the study (either as per protocol or prematurely terminated), actuarial analyses with the Kaplan-Meier method will be performed for each study outcome to illustrate effect differences. All analyses performed will be two-tailed and wondered as significant when p<0.05.

### Interim analyses and termination rules

This study will be terminated as per protocol when at least 43 patients complete 10 days of i.v. valproic acid therapy.

Premature termination will be executed when the following events occur:

- Since only a high effect size (>50%) in reducing death will be considered worthwhile to pursue further clinical study on valproic acid activity on COVID-19, the outcome rate boundary to terminate prematurely this proof-of-concept study will be 10 deaths occurred in ventilated patients.
- A rate of acute side effects (either thrombocytopenia, acute liver disease or acute pancreatic injury) exceeding 4 cases (10% event rate) will be considered unacceptable and will lead to premature termination, given that this rate exceeds the expected, given the current information on critically-ill patients exposed to valproic acid to treat epilepsy and status epilepticus.

## Conclusions

It is necessary for a useful drug to treat COVID-19 and its consequences. Given that it is expected the duration of a long-lasting pandemic,^25^ it can be supposed that we will have several ways to treat this disease, to improve several clinical outcomes of importance to eventually stop the direct sanitary implications and the very high economic burden that the COVId-19 pandemics has caused. Although long-term adverse effects and even pro-inflammatory consequences have been reported with the chronic use of VPA, given the urgent need for a drug active against COVID-19 to shorten the high mortality and LOS, the study of VPA is justified from a scientific standpoint.

## Data Availability

All data generated upon completion of this interventional study will be available in a proper repository.

## Acknowledgements

The authors are grateful to the authorities, colleagues, and patients of *Instituto Nacional de Ciencias Médicas y Nutrición Salvador Zubirán* for their support to perform this trial, as well as the grants of possible donors that are currently evaluating this investigation idea.

---

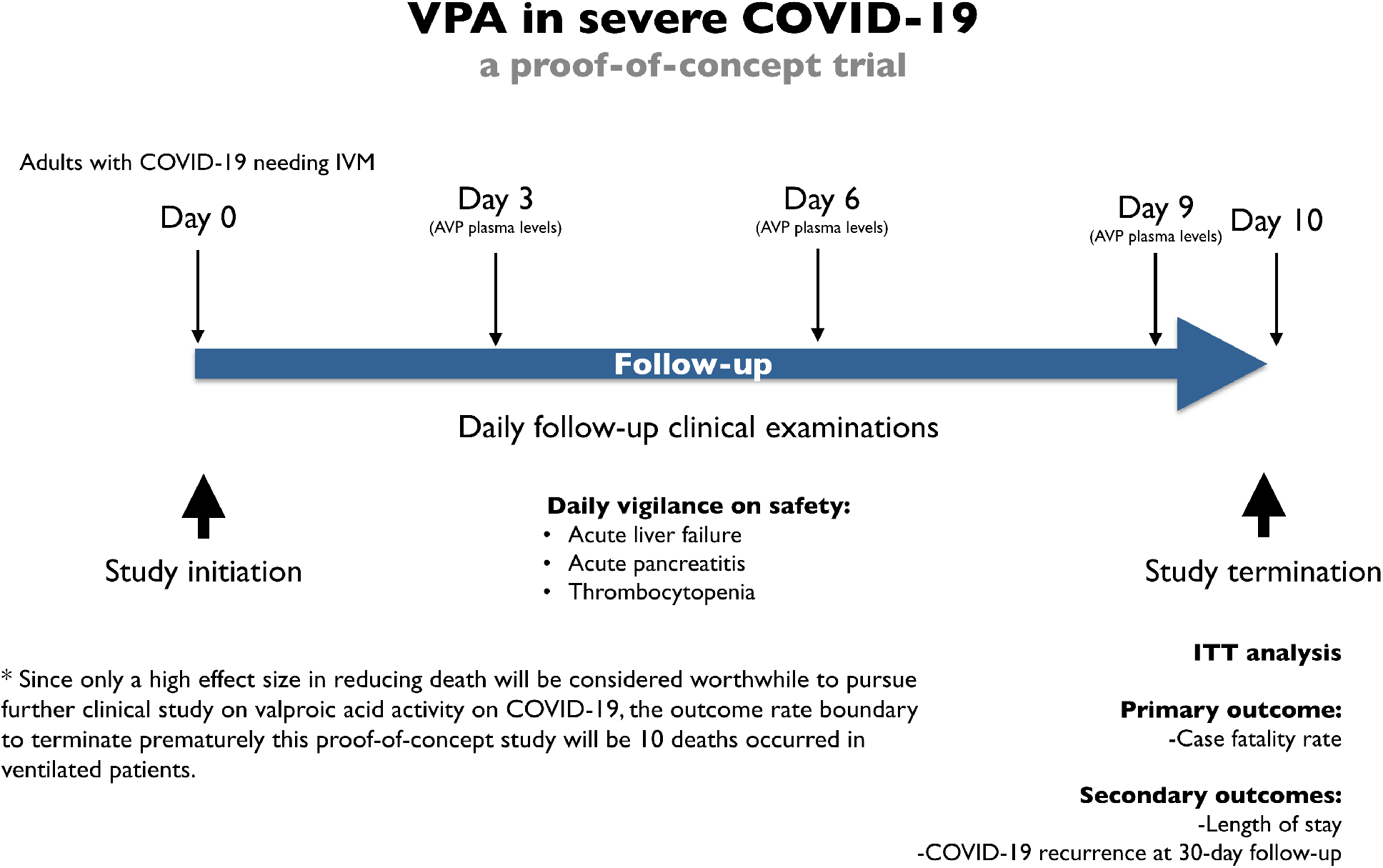
**Figure**. Diagrammatic study outline: methods of an open-label proof-of-concept trial of intravenous valproic acid for severe COVID-19.

